# Peripheral Immune Alterations and T Cell Clonal Dynamics Across the Parkinson’s Disease Spectrum

**DOI:** 10.64898/2026.09.23.26363800

**Authors:** Mikaela Rosen, J. Oriol Narcis Majos, Daniele Mattei, Elena Mejia, Carlos Perez Mandry, Beomjin Jang, Kailash BP, Tatsuhiko Naito, Amanda Allan, Tarek Khashan, Charlie Argyrou, Mengxi Yang, Deborah Raymond, Casey Young, Kelly Astudillo, Aloysius Domingo, Steven Frucht, Susan Bressman, Anna Tocheva, Giulietta Riboldi, Rachel Saunders-Pullman, Towfique Raj

## Abstract

The role of peripheral immunity in Parkinson’s disease (PD) remains incompletely understood. Here, we performed paired single-cell RNA sequencing (scRNA-seq) and T-cell receptor (TCR) profiling of 630k peripheral blood mononuclear cells (PBMCs) from 168 donors spanning idiopathic and genetic PD, prodromal PD, and healthy controls. We identify broad changes in immune-cell composition across the disease spectrum, including decreased naive B cells and increased naive CD8+ T cells in idiopathic PD. Transcriptional profiling reveals disease stage-dependent remodeling of mitochondrial and oxidative phosphorylation programs, with increased activity in prodromal disease followed by reduced activity in manifest PD, consistent with a transition in peripheral immune state during disease progression. High-resolution T cell analysis further identifies sex-dependent changes in cytotoxic CD8+ T-cell states and TCR clonal diversity. TCR motif analysis reveals disease-associated receptor groups with predicted specificities spanning PD-related, viral and autoimmune antigens. Together, these findings provide a comprehensive single-cell map of peripheral immune dysregulation in PD, defining dynamic and sex-dependent remodeling of peripheral immunity across the PD spectrum.

## Introduction

Parkinson’s disease (PD) is the second most common neurodegenerative disease, affecting more than 10 million people worldwide^1,2^. PD has a long prodromal phase before clinical diagnosis, during which substantial neuronal dysfunction and dopaminergic cell loss occur. By the time motor symptoms emerge, synaptic function is already markedly impaired and up to 60% of dopaminergic neurons may be lost^3^. Current treatments provide symptomatic benefit but do not prevent neurodegeneration, disease progression or conversion from prodromal to clinically manifest PD^4–6^. This highlights the need for biomarkers that capture early disease-associated biology and for therapeutic strategies that target mechanisms active before irreversible neuronal loss.

Although PD has traditionally been studied as a neuron-centered disorder, increasing genetic^7–9^, neuropathological^10,11^, and immunological^12,13^ evidence implicates systemic immunity in disease pathogenesis. Peripheral immune cells are attractive candidates for mechanistic and biomarker studies because they are accessible in blood and can reflect inflammatory state, antigen exposure, aging, genetic background, and disease-associated immune remodeling. However, the extent to which peripheral immune alterations in PD reflect changes in immune cell composition, transcriptional state or antigen-driven adaptive immune responses remains incompletely understood.

T lymphocytes (T cells) are of particular interest because they integrate inflammatory signaling, antigen specificity and human leukocyte antigen (HLA) -restricted immune recognition. Studies of circulating T-cell populations in PD have reported alterations in total, CD4+, CD8+, and regulatory T-cell compartments^14–18^, although findings have been inconsistent, likely reflecting limited cohort size, differences in disease stage, aging effects, genetic heterogeneity and the limited resolution of bulk or low-dimensional immune profiling. Beyond the blood, T-cell clonal expansion has been reported in the cerebrospinal fluid (CSF) of individuals with PD and CD4+ T cells have been detected near Lewy bodies and dopaminergic neurons in postmortem brain tissue^10,19^. Experimental models further support a functional role for T cells, with regulatory T cells showing protective effects and CD4+ T cells contributing to neurodegeneration in PD models ^11,20–22^.

The genetic architecture of PD further supports a role for adaptive immunity. Genome-wide association studies have identified 78 PD-risk loci^9^, including variants within the human leukocyte antigen region^23,24^, suggesting that antigen presentation and HLA-restricted T-cell responses may contribute to disease susceptibility or progression. Consistent with this possibility, peripheral T cells reactive for α-synuclein have been detected in individuals with PD^25,26^ and associated with diagnosis, disease duration, and clinical severity^25–28^. These findings suggest that peripheral T-cell responses may not simply reflect downstream inflammation, but could encode disease-relevant immune recognition and provide insight into patient stratification and therapeutic targeting.

Despite this growing evidence, the peripheral immune landscape of PD remains poorly resolved. Most studies have focused on selected immune populations, bulk inflammatory markers, or predefined antigen-specific responses, limiting the ability to distinguish broad immune remodeling from disease-associated transcriptional programs or clonally expanded T-cell responses. In addition, aging and sex are major drivers of immune composition and function and must be considered when interpreting immune changes in neurodegenerative disease^29–31^. As a result, it remains unclear how aging, sex, genetic predisposition, disease status, immune cell composition, pathway-level dysregulation, T-cell subpopulation structure, clonal expansion and antigen specificity interact across the PD spectrum.

Here, we performed single-cell transcriptome and paired T-cell receptor sequencing of peripheral blood mononuclear cells from a PD-focused cohort spanning controls and disease-relevant clinical groups. We first characterized the immune cell populations present in peripheral blood and assessed compositional changes associated with aging, sex and disease. We then defined pathway-level transcriptional immune dysregulation across the PD spectrum and examined whether T-cell subpopulation structure was associated with disease-related clonal expansion. Finally, we investigated clonally expanded T cells and used antigen-specificity prediction to highlight potential candidate antigenic epitopes. Together, this study provides an integrated single-cell and TCR-resolved map of peripheral immune dysregulation in PD and highlights adaptive immune features with potential relevance for biomarkers, disease mechanisms, and therapeutic development.

## Results

### Single-cell transcriptome and TCR sequencing of PBMCs in PD-focused cohort

We performed a comprehensive single-cell RNA-seq (scRNA-seq) analysis of human peripheral blood mononuclear cells (PBMCs) with paired single-cell V(D)J T-cell receptor (TCR) immune repertoire sequencing (**Figure 1A**). The cross-sectional cohort comprised donors with genetic and idiopathic forms of PD (n=91), prodromal cases who are at elevated risk of developing PD with diagnosed Rapid Eye Movement (REM) sleep behavior disorder (RBD; n=20), non-manifesting carriers of PD-associated *GBA1* or *LRRK2* mutations (n=10) and controls with no known neurological disease (n=47; **Table 1**). Donors were recruited from the The Marlene and Paolo Fresco Institute for Parkinson’s and Movement Disorders at the New York University (NYU) Langone Health and The Mount Sinai-Beth Israel Movement Disorders Clinic at the Mount Sinai Hospital (MSH). Donors were age-and sex-matched across diagnoses with a mean age of 68.35 (range 49-87) and had approximately balanced sex ratios (**Table 1** and **Supplementary Table 1**). As both PD and RBD are more prevalent in males, a slight male enrichment is present in the data^32,33^. PBMCs were processed promptly after blood draws and cryopreserved under standardized conditions to minimize technical batch effect. From 176 initial samples, we sequenced over 980k cells. After rigorous quality control (QC) (see Methods, **Figure S1** and **Supplementary Table 2**), we maintained robust transcriptome data for 168 unique donors and over 630k cells (**Figure 1B**). T cells constituted an average of 61% of PBMCs (**Figure 1C**), yielding transcriptome data for 363k T cells with paired TCR data from 337k (approximately 93%; **Figure 4B**).

**Figure 1.**
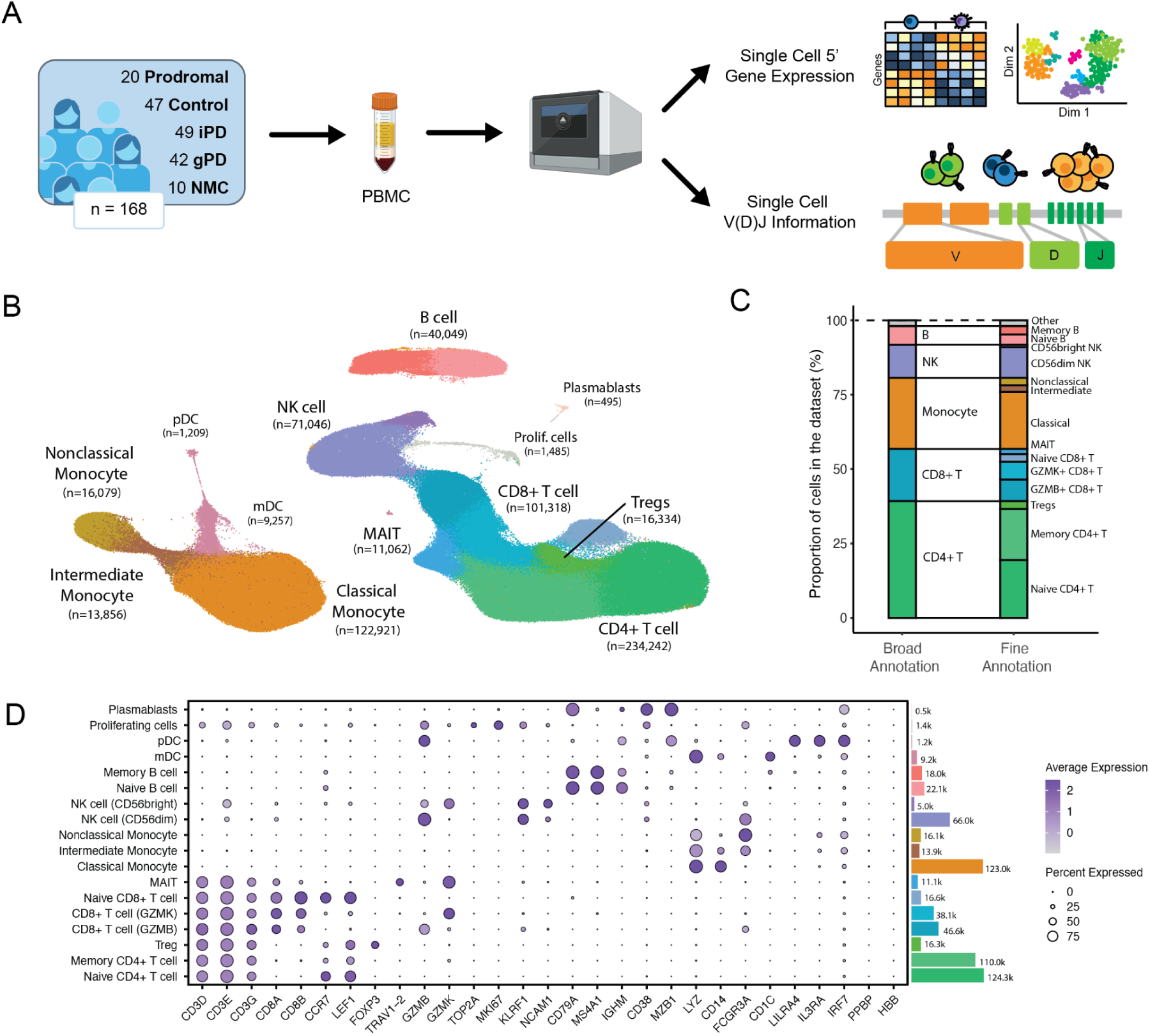
Cohort characterization and immune cell type identification in peripheral blood mononuclear cells (PBMCs). **A)** Schematic overview of the study design illustrating PBMC collection and parallel generation of single-cell 5’ gene expression and V(D)J libraries. **B)** Uniform Manifold Approximation and Projection (UMAP) of 630k PBMCs across 168 donors after quality control (∼4,000 cells per donor), colored by cell type annotation. **C)** Stacked bar plots showing cell type proportions at broad (8 clusters) and fine (18 clusters) annotation levels, colored as in B. **D)** Dot plot of representative marker gene expression across fine-annotated cell types. Dot size reflects the percentage of cells expressing each gene and color intensity reflects log-normalized average expression. Bars on the right indicate the total number of cells per cluster, colored as in B and C. Prodromal are patients with REM Sleep Behavior disorder; iPD is idiopathic PD; gPD is genetic PD including patients with either or both *GBA1* and *LRRK2* mutations. NMC are non-manifesting carriers who have either *GBA1* or *LRRK2* mutations but no PD diagnosis.

**Table 1.** Demographic and clinical characteristics of the study cohort. Participant demographics are shown for healthy controls (n = 47), idiopathic PD (iPD, n = 49), *GBA1* variant-associated PD (*GBA1-*PD, n = 24), *LRRK2* variant-associated PD (*LRRK2*-PD, n = 15), non-manifesting carriers of genetic mutations (NMC, n = 10) and REM sleep behavior disorder (RBD, n = 20). Age at recruitment is reported as mean (standard deviation). Sex, self-reported ethnicity, and predicted genetic ancestry are reported as counts and percentages. Three donors who are diagnosed with PD and have both *GBA1* and *LRRK2* mutations are included in our final 168-donor cohort but are not shown in this table.

|  | Controls |  |  | PD |  |  | Prodromal |
| --- | --- | --- | --- | --- | --- | --- | --- |
|  | Control | NMC *GBA1* | NMC *LRRK2* | iPD | *GBA1*-PD | *LRRK2*-PD | RBD |
| Total Donors | 47 | 6 | 4 | 49 | 24 | 15 | 20 |
| Age of Onset | *NA* | *NA* | *NA* | 62.1 (8.3) | 56.6 (10.1) | 64.5 (11.1) | *NA* |
| Age at Recruitment | 69.2 (6.9) | 64.8 (5.6) | 65.8 (5.4) | 68.6 (7.1) | 65.3 (8.3) | 74.9 (8.4) | 66.5 (4.6) |
| Sex |  |  |  |  |  |  |  |
| F | 26 (55%) | 3 (50%) | 2 (50%) | 21 (43%) | 10 (42%) | 7 (47%) | 5 (25%) |
| M | 21 (45%) | 3 (50%) | 2 (50%) | 28 (57%) | 14 (58%) | 8 (53%) | 15 (75%) |
| Reported European Ethnicity | 43 (91%) | 6 (100%) | 4 (100%) | 42 (86%) | 15 (63%) | 15 (100%) | 18 (90%) |
| Predicted AJ Ancestry | 25 (53%) | 6 (100%) | 4 (100%) | 30 (61%) | 23 (96%) | 15 (100%) | 6 (30%) |

Following cluster-based cell type annotation and manual marker curation (**Supplementary Table 3**), we annotated 8 major PBMC cell types that were further divided into 18 fine annotations (**Figure 1C** and **Figure S2**). T cells were identified by CD3 receptor gene expression (*CD3D*, *E* and *G*), with *CCR7* and *LEF1* distinguishing naive T cells from more cytotoxic T cells that express granzyme genes *GZMB* or *GZMK*. NK cells express granzymes but they also show expression of *KLRF1* and *NCAM1* (CD56), with the degree of expression distinguishing subpopulations. More specific subtypes of T cells were also identified such as T regulatory helper cells (Tregs) which show a *FOXP3* gene signature and mucosal associated invariant T cells (MAIT) with the distinctive expression of *TRAV1-2* T-cell receptor gene. B cells expressed *CD79* and *MS4A1* (CD20). Monocytes cluster with strong *LYZ* expression and varying *CD14* and *FCGR3A* (CD16) expression, with classical monocytes having the strongest expression of the former and nonclassical monocytes having the strongest expression of the latter. Cells from the major immune lineages (T cells, B cells, NK cells, and monocytes) were represented in all donors at the expected proportions (**Supplementary Table 4**). We also identified rare populations of proliferating cells (*TOP2A*, *MKI67*) and plasmablasts (*MZB1*, *CD38*), along with a small population expressing platelet-associated markers such as *PPBP* (**Figure S1**; see Methods). These underrepresented and non-target populations were excluded from downstream analyses.

### PBMC composition is shaped by aging, sex, and disease status

To assess cell type compositional changes across biological covariates, we applied two complementary approaches: Covarying Neighbor Analysis (CNA), which identifies transcriptional neighborhoods enriched across disease stages (**Figure 2A**), and linear regression model (crumblr^34^) to confirm findings and explore interaction effects (**Figure 2B**). Despite their fundamentally different frameworks, the two approaches yielded highly concordant results (**Figure S3A** and **Supplementary Table 5**).

**Figure 2.**
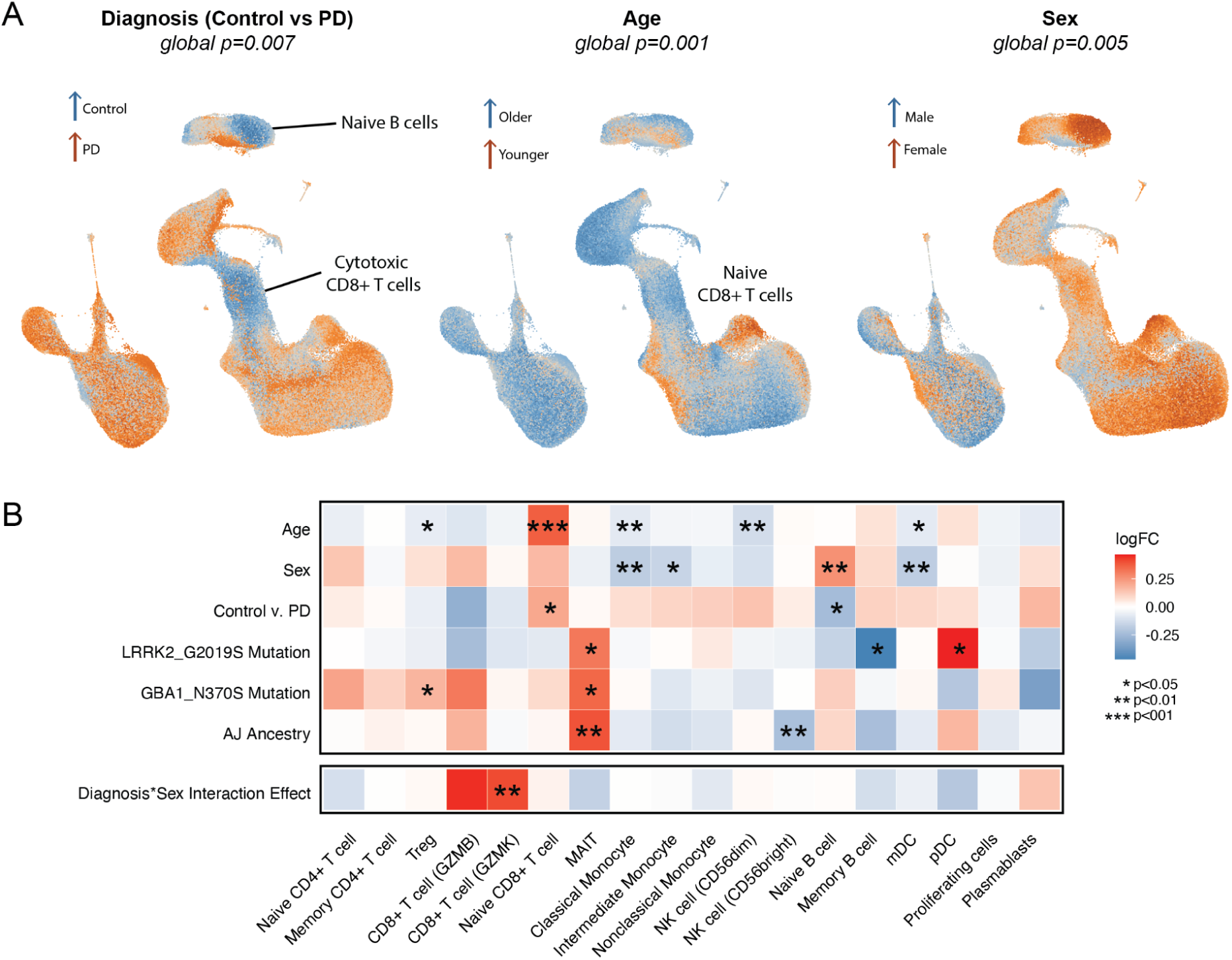
Cell type composition changes at the whole PBMC level in PD. **A**) UMAP of PBMCs colored by the correlation of each cell’s transcriptional neighborhood with three biological covariates: diagnosis (Control and NMCs vs. PD), age, and sex (left to right). Cool colors (blue) indicate enrichment in controls, older donors, and males, respectively; warm colors (orange) indicate enrichment in PD, younger donors, and females. CNA global p-values are shown above each UMAP. Annotated populations highlight the cell types driving the strongest associations. **B**) Heatmap of log fold-change estimates from a linear regression model (crumblr) across fine-annotated PBMC cell types for each covariate, including diagnosis, age, sex, genetic covariates (*LRRK2* G2019S, *GBA1* N370S, Ashkenazi Jewish ancestry), and a diagnosis by sex interaction term. Color intensity reflects the magnitude of the log fold-change; asterisks denote statistical significance (*p < 0.05, **p < 0.01, ***p < 0.001). Warm colors indicate enrichment in PD, younger donors, females, the presence of G2019S or N370S mutations and AJ ancestry.

The strongest driver of compositional variation was age (CNA global p = 0.001), even within the relatively narrow age range represented within our cohort (49-87 years). Naive CD8+ T cells were significantly depleted in older donors (p < 0.001; **Figure 2A-B**), consistent with the well-established contraction of the naive T-cell compartment with immunosenescence. Aging was further associated with shifts in some scattered T cell subsets, Tregs, and classical monocytes (**Figure 2B**). Sex was an independent compositional driver (CNA global p = 0.005) as well (**Figure 2A**). Males showed higher proportions of classical and intermediate monocytes, consistent with prior reports of male-biased myeloid frequencies in peripheral blood^29,35–37^ and lower naive B cells compared to females (**Figure 2B**). Ashkenazi Jewish (AJ) ancestry was additionally associated with differences in MAIT and NK cell proportions (**Figure 2B**), reflecting the genetically distinct composition of this subgroup within our cohort.

Diagnosis-associated compositional shifts were observed at CNA global p = 0.007 (**Figure 2A**). Naive B cells were more abundant in control blood (p < 0.05; **Figure 2B**), consistent with previously reported B cell depletion in PD^38^. Conversely, naive CD8+ T cells were elevated in iPD (p < 0.05; **Figure 2A-B**), accompanied by a corresponding reduction in cytotoxic CD8+ T-cells, significant in the neighborhood-based CNA analysis, suggesting an overall shift toward a less cytotoxic peripheral T-cell state in PD blood (**Figure 2A**). *GBA1* N370S and *LRRK2* G2019S mutation carriers showed distinct compositional profiles, with *GBA1* mutation showing altered Treg and MAIT cell proportions and *LRRK2* mutation associated with shifts in Memory B cells and pDC subsets (**Figure 2B**).

To explore sexual dimorphism in disease-associated immune remodeling, we applied a linear regression interaction model using centered log-ratio (CLR)-transformed cell type counts, sharing all covariates with the main effects model; results from the two models correlated well (**Figure S3 C-D**). A significant diagnosis by sex interaction was identified for cytotoxic CD8+ GZMK+ T cells (p < 0.01; **Figure 2B**). Notably, the directionality of this association was sex-dependent. Cytotoxic CD8+ GZMK+ cells were more abundant in female PD blood relative to female controls, whereas the opposite pattern was observed in males (**Figure 2B**). This sex-specific divergence in cytotoxic T cell composition highlights the importance of incorporating sex as a biological variable in PD immune profiling.

### Transcriptional Immune Profiling Across the Parkinson’s Disease Spectrum

To characterize transcriptional differences across disease states, we performed differential gene expression (DGE) analysis comparing the transcriptomes of control, prodromal RBD and iPD. We applied a pseudobulk framework using DESeq2, restricting analyses within each cell type to genes with sufficient expression (3,000-10,000 genes) and excluding outlier samples to minimize technical bias (see Methods; **Figure 3A**; **Figure S4 B-D**). Consistent with prior reports showing increased transcriptional changes in Prodromal cases with higher phenoconversion risk^39^, our analysis revealed more pronounced transcriptomic changes in Prodromal relative to both controls and iPD, whereas differences between controls and iPD were more modest (**Figure 3A** and **Supplementary Table 6**). When comparing the RBD prodromal group to either healthy controls or iPD, the median absolute fold change is 0.1. In contrast, the difference between iPD and controls is much smaller, with a median fold change of only 0.03. We detected deregulated genes across all cell types and diagnosis comparisons using a nominal significance cutoff of p<0.01 and across most cell types when comparing to prodromal using an adjusted cutoff FDR <0.05 (**Figure 3A**). When comparing control and iPD PBMCs, there are a few genes deregulated at an adjusted significance threshold.

**Figure 3.**
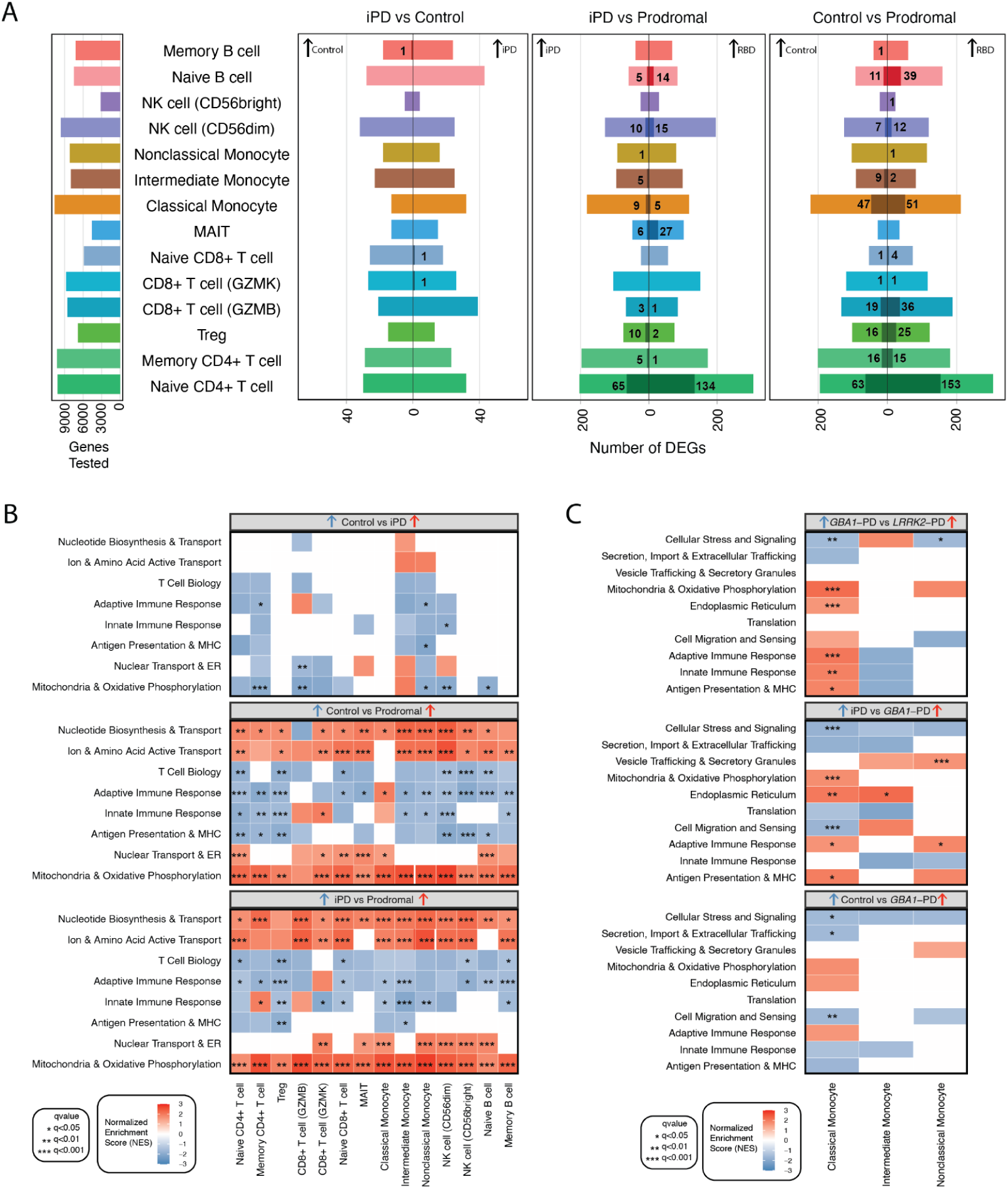
Differential expression and pathway enrichment across PBMC populations in idiopathic, prodromal and genetic PD. **A)** The number of genes tested and of differentially expressed genes (DEG) from pseudobulked DESeq2 analysis using nominal p value threshold<0.01 (outer bar) and adjusted p-value cutoff FDR<0.05 (inner shaded bar) comparing iPD, Control and Prodromal PBMCs. The number of DEGs meeting FDR<0.05 are indicated with the shaded inner bars and numbers. **B)** Most significant pathways enriched among expressed genes in pairwise comparison of control, RBD prodromal and iPD. **C)** Most significant pathways enriched among differentially expressed genes comparing *GBA1-*PD to iPD, *LRRK2*-PD and controls. Input to GSEA was sorted by signed log fold change DESeq2 values.

To characterize the biological processes underlying immune cell transcriptional changes across the PD disease spectrum, we performed gene set enrichment analysis (GSEA) on the pseudobulk differential expression results across six pairwise comparisons: iPD vs Control, Prodromal vs Control, Prodromal vs iPD, *GBA1-*PD vs Control, *GBA1-*PD vs iPD and *GBA1-*PD vs *LRRK2*-PD (**Supplementary Table 7**). Enrichment was assessed across all major peripheral blood immune cell types, including naive and memory CD4+ and CD8+ T cells, regulatory T cells (Tregs), MAIT cells, classical, intermediate, and nonclassical monocytes, NK cell subsets (CD56dim and CD56bright), and naive and memory B cells. The most consistent and cross-cutting finding across all six comparisons was the dysregulation of mitochondria-related pathways (q<0.05-0.001 across cell types and comparisons). Mitochondria & Oxidative Phosphorylation was significantly enriched across multiple immune cell types and comparisons, including upregulation in widespread cell types in the Prodromal subpopulation compared to the iPDs and to Controls (q<0.001 in almost all cell types). Strikingly, the same pathways were downregulated in iPD when compared to Controls **(Figure 3B)** suggesting that compensatory mitochondrial upregulation is an early and detectable immune transcriptional change in PD, predating clinical diagnosis. Beyond mitochondrial dysregulation, the Prodromal vs. Control comparison revealed the broadest transcriptional changes overall, we observe downregulated pathways related to both innate and adaptive immune response in the Prodromal group. On the other hand, pathways related to cell biosynthesis, homeostasis and molecular transport were positively enriched in the same population, suggesting an active shift in cellular metabolic and homeostatic programs at the prodromal stage **(Figure 3B).** The same trend was observed in the Prodromal vs iPD comparison.

Next, building on our previous and extensive research on monocyte characterization in the context of PD^12,40,41^ we sought to further investigate, at single-cell resolution, the effects of *GBA1* and *LRRK2* mutations across the three major monocyte subtypes. In the *LRRK2*-PD vs. *GBA1-*PD comparison, Classical Monocytes exhibited significant enrichment of Mitochondria & Oxidative Phosphorylation, Endoplasmic Reticulum, Adaptive and Innate Immune Response pathways in *LRRK2*-PD, while Nonclassical Monocytes showed enrichment of Cellular Stress and Signaling in *GBA1-*PD, suggesting divergent monocyte subset responses across genetic subtypes. The *GBA1-*PD vs. iPD comparison yielded the broadest transcriptional differences, with Classical Monocytes displaying significant enrichment in *GBA1-*PD of Cellular Stress and Signaling (q<0.001), Mitochondria & Oxidative Phosphorylation (q<0.001), Endoplasmic Reticulum (q<0.01), Cell Migration and Sensing (q<0.001), and immune effector pathways (q<0.05), while Nonclassical Monocytes exhibited upregulation of Vesicle Trafficking & Secretory Granules (q<0.001). In contrast, the *GBA1-*PD vs. Control comparison revealed comparatively limited transcriptional differences, restricted largely to Cellular Stress and Signaling (q<0.05), Secretion Import & Extracellular Trafficking (q<0.05), and Cell Migration and Sensing in Classical Monocytes (q<0.01). Taken together, this suggests that *GBA1-*PD monocytes are transcriptionally more divergent from iPD than from healthy controls **(Figure 3B)**.

### High-Resolution T Cell Annotation Identifies Genotype-and Sex-Dependent Shifts in PD Blood

More than half of the PBMC dataset consists of T cells, providing sufficient resolution to interrogate T-cell subpopulation structure. After subsetting and reclustering T and NK cells, the seven T-cell fine annotations (**Figure 1C**) were even more specifically characterised into eighteen subpopulations using reference based mapping^42,43^ and marker genes (**Figure 1A** and **Supplementary Table 3**) and validated against the previously published profiles described in Monaco Immune^44^ (**Figure S5C**). Naive T cells are most abundant in CD4+ with over 100k cells sequenced which equates to an average of 28% of total CD4+ T cells. On the other hand, naive T cells are only on average 4% of the CD8+ T cell population with the most abundant subtype being GZMB+ (**Figure 4B** and **Supplementary Table 8**).

CD4+ memory T cells were annotated further and split into seven subtypes (**Figure 4A**). The Treg cluster, marked by both *FOXP3* and *IL2RA* expression, remained robust and was not reannotated. Follicular helper (Tfh) cells showed stronger relative expression of *ICOS* and *CXCR5*. Th1 cells showed a gene expression signature including high *CXCR3*, *CCL5* (RANTES) and low *CCR6* **(Figure S2B**). In contrast, cells in the Th17 annotated cluster expressed low *CXCR3* and high *CCR6*. The Th2 cluster was characterized by its expression of *CCR4* and *GATA3* with a subset of cells also expressing *KLRB1* earning the further annotation of activated (**Figure S2B**). However, because of the small size of the activated population, this annotation was collapsed back into a unified Th2 cluster (**Figure 4A**). We also identified another more activated CD4+ population which we call CD4_KLRB1 and is characterized by high *KLRB1* expression. The most cytotoxic cluster of CD4+ T cells was annotated as CD4+ cytotoxic T lymphocytes (CTL) and is characterized by the strong CD4 expression accompanied by the expression of several cytotoxic genes including *GZMA*, *GZMH*, *CST7* and *KLRB1*.

CD8+ T cells were segregated further from two major populations into six more specific groups (**Figure 4A**). MAIT cells remain clustered the same as previous, marked by *TRAV1-2* and *SLC4A10*. The GZMB+ and GZMK+ populations that were identified at the whole PBMC annotation level remain robust clusters marked by the expression of their namesake genes. The more abundant GZMK+ cells express cytotoxic genes *GZMA* and *NKG7*. Whereas the more cytotoxic GZMB+ expresses *GZMH*, *FGFBP2* and *GNLY* (**Figure 4B**). The KLRF1+ and KLRC1+ groups express a somewhat combined profile compared to the other two aforementioned cytotoxic groups and have strong expression of their namesake genes. The terminally differentiated effector memory T cells (TEMRA) have a strong cytotoxic profile and *TOX* expression. Upon reclustering, the gdT cells can be parsed out in the low dimensional representation. This cluster has a generally cytotoxic expression signature which is similar to other cell types but it also specifically shows expression of T cell receptor gamma genes such as *TRGC1*. The best way to further confirm its identity is by exploring our V(D)J library since we only sequenced alpha-beta and not gamma-delta receptors. This cluster has little to no mapping of the V(D)J (**Figure 4A and B**).

Several of the key alterations found in cell type composition at the whole PBMC level involved the T cell populations. Therefore, we re-examined the cell type composition differences within the T-cell compartment using our more specific clustering and annotations. Again, we apply both a cell type agnostic and cell type sensitive approach to explore the cell type proportion changes (see Methods). Many of the major results recapitulate PBMC level analysis **(Figure 2 A-B**; **Figure 4C)**. For example, the strongest association is once again between differential cell abundance of Naive CD8+ T cells and advanced age **(Figure 4 C-D; Figure S6A)**. Similarly, proportions of Memory CD4+ T cells and different helper T cell subtypes (Tfh, Th2 and Tregs) are also associated with advanced age **(Figure 4C)**. We observe a significant relative increase in Th2 cells in males in both control and iPD blood **(Figure 4C, E)**. Another finding that is confirmed at the T-cell clustering level is the main effect for more naive CD8+ T cells in PD and the trend towards less cytotoxic cell types in PD (**Figure 4C**). The interaction effect found between Diagnosis and Sex for GZMB+ cells at the PBMC level is now more specifically attributed to the CD8+ KLRF1 and TEMRA cells which have clustered out from the GZMB+ (**Figure 4C**). CD8+ KLRF1 T cells and TEMRA cells are more abundant in males in Control, but are observed in higher proportions in female PD (**Figure 4F-G**). If you explore this interaction further, you can notice that there is also a change in the detected clonal expansion sizes, if we observe more cells we also capture more cells belonging to large clonal expansions (**Figure 4 F-G**). This indicates that changes in composition may be related to clonal expansions we observe.

### Clonally expanded T cells in the blood

After characterizing the cell subtypes present within the T cell compartment and observing alterations of the cytotoxic T cell subtypes, we sought to describe the clonal attributes of these clusters. Clonal expansion was defined as groups of T cells with identical complementary-determining region 3 (CDR3) nucleotide sequences in both alpha and beta chain of their TCR. As expected, we found more clonal expansion within the CD8+ compared to the CD4+ T cell compartment (**Figure 4A and B**). The most clonally expanded cell states were the CD8+ GZMK/B+ memory, CD4+ CTLs and TEMRA cells (**Figure 4B**). These cell types are also considered the most cytotoxic cell types (**Figure S5B**). Clones larger than 20 cells were predominantly composed of CD8+ GZMB+ cells. Cells from the top 50 clones revealed striking differences in distribution across cell types (**Supplementary Table 9**). The largest two clones with over 600 cells are both dominated by TEMRA cells **(Figure 5 A-B)**. Other hyperexpanded clones are more polarized to the cytotoxic CD8+ T cell subtypes, including CD8+ GZMK+ and CD8+ GZMB+ **(Figure 5 A-B, clones b and d)**. There are also clones dominated by cytotoxic CD4+ T cell subtypes such as CD4+ CTL and Th1 cells **(Figure 5 A-B, clones c and e)**.

**Figure 4.**
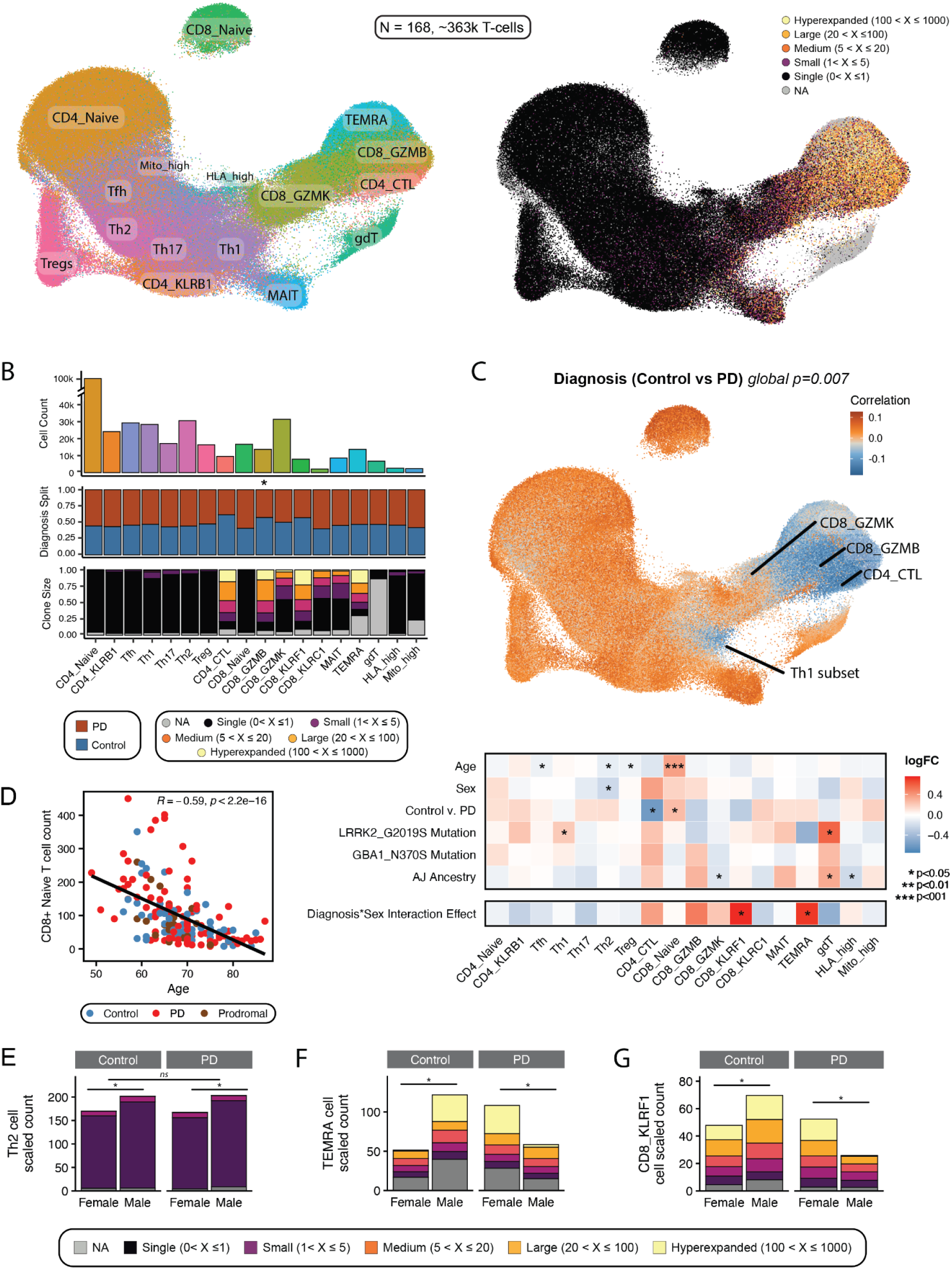
T cell composition changes are related to differences in clonal expansion. **A)** UMAP of T cells colored by cell type (left) and clonal expansion (right). **B)** Stacked barplot showing total T cell counts, proportion of cells from each diagnosis and clonal expansion per cell type. Asterisks denote changes meeting p<0.05 significance level as determined with a two sided T-test. **C)** UMAP of T cells colored according to their neighborhood’s abundance correlation to diagnosis. CNA global p-value is shown. The lower section shows Crumblr cell type composition results using an interaction model. Asterisks denote changes meeting various significance level thresholds. **D)** Cell type counts for Naive CD8+ T cells correlated with age, nominal p-value significance shown. **E-G)** Cell type counts for various T cell subtypes, Th2 cells **(E)** TEMRA **(F)** and CD8_KLRF1 **(G)**, filled with proportion of clonal expansion (shared legend below).

**Figure 5.**
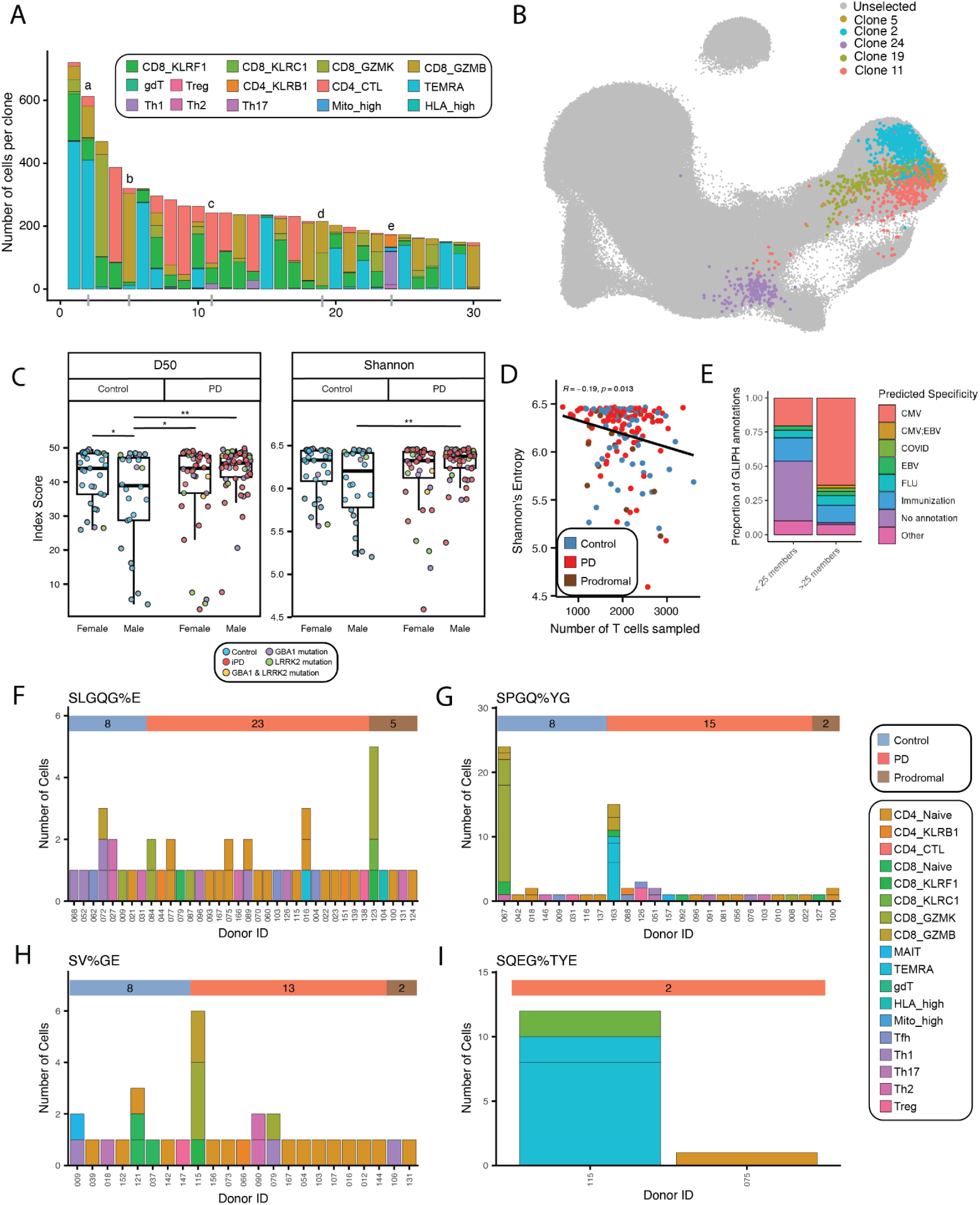
T cell clonal expansions, repertoire diversity and antigen specificity prediction. **A)** Barplot showing the top 30 largest clonal expansions present in our cohort and their cell type composition. Selected clones are indicated a-e as representatives of differently composed clonotypes. **B)** T cell UMAP with selected clones highlighted and colored by their dominant cell type. **C)** Diversity metrics index scores D50 and Shannon’s entropy split by diagnosis and sex; asterisks denote nominal statistical significance (*p < 0.05, **p < 0.01). **D)** Correlation plot between the number of T cells sampled per donor and Shannon’s diversity index. Spearman’s correlation was used to test strength and significance, nominal p-value significance shown. **E)** Proportion of reference predicted antigen specificity annotations in shared motif GLIPH analysis split by number of distinct members in each group. Predicted specificities were simplified down based on strength of evidence and favoring more common viral antigens. **F-I)** PD-enriched GLIPH motif groups with annotations including PD antigens. Number of cells with motif-matching CDR3b per donor grouped by diagnoses. The top grouping bar indicates the number of donors in the group from each diagnosis. All clones are dominated by PD and prodromal cases. Shared legend on the right.

Beyond exploring specific clones and the raw size of clonal expansions, T cell diversity metrics can summarize the level of clonal expansion across a TCR repertoire (**Supplementary Table 10**). T cell diversity was measured by both D50 index and Shannon’s entropy. The D50 index focuses on repertoire dominance by the most frequent clones, with larger values indicating less dominance by the top clone and hence more diversity. Shannon’s entropy highlights the number of distinct clonotypes (richness) and the clone size distribution (evenness), with larger values indicating more distinct clonotypes and evenly distributed clone sizes which also means there is more diversity in the repertoire. There are no statistically significant differences in clonal diversity metrics attributed to diagnosis (p=0.08). However, there is a statistically significant difference driven by both sex and diagnosis, where male control donors show lower D50 indices and therefore more clonal expansion compared to female control blood (p<0.01) and compared to both sexes in PD blood (p<0.05 for females, p<0.01 for males) **(Figure 5C)**. The observed difference is more significant when comparing D50 indices as opposed to Shannon’s entropy. It is also worth noting that the diversity metrics are affected by the number of cells, and most importantly, the number of T cells that are sampled. There is an inverse correlation between Shannon’s entropy and the number of cells sampled (r=-0.19, p<0.05; **Figure 5D**), wherein sampling more cells results in greater capacity to capture T cells with the same TCR and that are from the same clone.

#### Antigen specific T cells and candidate antigenic epitopes

Having established which T-cell states are clonally expanded and how repertoire diversity varies with sex and diagnosis, we next asked what these expanded clones might recognize. We predicted specificities using GLIPH and a compiled reference database of previously reported epitopes and CDR3 pairs^45–49^. Exact matching of β-chain CDR3 sequences with those from reference databases yielded 38,269 matches (∼10% of our T cells), distributed across cell types and drawn predominantly from single or small clones.

Since shared motifs capture antigen specificity more broadly than exact matches, we ran GLIPH on our CDR3s together with the reference database, yielding 167,979 groups; 63,278 remained after requiring at least three samples and three amino acids per motif and contained both cohort and reference sequences (**Supplementary Table 11**). Taking the dominant reference annotation per group, cytomegalovirus (CMV) was the most frequent predicted specificity, particularly in groups spanning more than 25 donors (**Figure 5E**). As with the exact matches, these groups consisted mostly of non-expanded clones, and few hyperexpanded clones carried reference annotations: the second largest clone (>600 cells; **Figure 5A-B, clone a**) fell in motif group %PDRGTE, annotated for HTLV-1, and the fifth largest (**clone b**) in S%QGNYG, annotated for an influenza epitope.

We then focused on the 336 GLIPH groups containing PD-specific reference CDR3s, of which 64 passed a more stringent Fisher’s cutoff (p<0.05) and 33 were enriched for PD and RBD prodromal donors (**Supplementary Table 12**). SLGQG%E (annotated for PD, CMV and autoimmune epitopes) contains an RBD prodromal donor with five expanded cells of largely cytotoxic phenotype (CD8+ KLRC1+ and GZMK+) plus one naive CD8+ T cell sharing the same TCR, suggesting a common lineage. Two PD donors contribute GZMK+ and TEMRA cells, whereas cells from control donors are skewed towards Th1 and Th2. SPGQ%YG, enriched 2:1 for PD and prodromal donors and annotated for two PD epitopes along with immunization and flu, shows expansion in four donors spanning *GBA1-*PD, *LRRK2*-PD and iPD, dominated by TEMRA, Th1/Th17 and Treg/Tfh cells. The only large control expansion is instead composed of cytotoxic CD8+ GZMK+ and GZMB+ cells (**Figure 5G**). The same PD-versus-control contrast in cellular composition holds for SV%GE (PD, dengue and CMV annotations; **Figure 5H**). Finally, SQEG%TYE matches a single reference antigen, a PD epitope, and contains only PD donors, with expansion of TEMRA and KLRF1+ cells in a donor that is also expanded within SV%GE (**Figure 5I**).

## Discussion

Given the role of inflammatory processes from the brain through to the periphery in PD, thorough study of the immune compartment is essential. Postmortem studies report lymphocyte infiltration in the brain^10^, increased levels of activated T cells in CSF^19^ and the presence of alpha-synuclein reactive T cells in patient blood particularly in early years of PD^25–27^. Our study, leveraging paired single-cell RNA and TCR sequencing, provides a detailed assessment of the relationship between the immune repertoire and cell state composition in a PD-focused cohort.

Pathway-level characterization of peripheral immune transcription reveals mitochondrial dysfunction, which has long been recognized as a central feature of PD neurodegeneration, from Complex I deficiency in substantia nigra^50,51^ and PD-linked genes *PINK1*, *Parkin*, and *DJ-1*, all of which regulate mitochondrial quality control^52,53^. Our data extend this concept to the peripheral immune compartment, demonstrating that mitochondrial transcriptional dysregulation is not confined to neurons but is a system-wide feature detectable in circulating monocytes, T cells, and NK cells.

Whereas previous studies^12,40,41^ reported downregulation of mitochondrial-related pathways in monocytes from individuals with PD, by expanding the analysis across multiple immune cell types and including prodromal subjects, our study reveals a more dynamic pattern. Mitochondrial pathways are upregulated in prodromal PD relative to controls but downregulated in iPD compared to controls. This biphasic trajectory, characterized by an early compensatory phase followed by later mitochondrial transcriptional collapse. Whereas prior studies^12,54^ primarily captured the collapsed state in manifest disease, our data provide evidence for a preceding compensatory immune-cell phase, with important implications for defining therapeutic windows and developing stage-specific biomarkers.

Prodromal immune signatures suggest a trajectory of progressive activation. Our observation of a predominantly upregulatory immune signature in the prodromal stage, spanning mitochondrial, metabolic, nuclear transport, and innate immune pathways is consistent with signatures reported by recent work from Zhang and colleagues^39^. In iPD, this shifts toward a mixed pattern, suggesting an evolving immune response that resembles immune exhaustion in chronic inflammatory states and may reflect sustained engagement with accumulating α-synuclein pathology. The early engagement of B cells and adaptive immune pathways at the prodromal stage is an intriguing finding that warrants further investigation, as autoantibodies against alpha-synuclein and other PD-related antigens have been reported in patient blood and may precede motor symptom onset^55,56^.

Previous studies have reported alterations in blood cell type composition, particularly within the T cell compartment^15,18,30^, but there are disparities likely due to limitations, including sample size and uncontrolled biological heterogeneity. We report an inverse association with naive CD8+ T cells with aging^29,57^ and identify iPD-associated decreases in naive B cells consistent with a prior smaller-scale scRNA-seq study^38^. Notably, naive CD8+ T cells are increased in PD, counter to their aging-associated decrease, suggesting disease-specific immune remodeling rather than an aging artifact. Grandke et al. 2025^30^ reported a similar increase in CD8+ T cells in a cohort of neurodegenerative patients, but only in female donors. Our work suggests that these changes are in fact disease related and not sex-specific effects.

Both aging and sex are major drivers of immune composition and must be considered when interpreting immune changes in neurodegenerative disease^29–31^. PD exhibits distinct sexual dimorphism, with a male-to-female ratio of approximately 1.5 that increases with age^32,58^. Even a recent immune focused clinical trial (Azathioprine) has reported that clinical effects could be different and greater in females than males^59^. A sex-dependent differentiated peripheral inflammatory response may, in part, be related to differences in PD presentation in males and females.

Our interaction analysis to explore sex-specific differences in cell type composition in PD identified that cytotoxic T-cell populations change in a sex-specific manner. CD8+ KLRF1 and TEMRA cells (more broadly attributed to cytotoxic CD8+ GZMK+ T cells) were higher in male compared to female controls but female compared to male PD blood. Increased non-memory CD8+ T cells have been observed in female PD patients^30^, though differential annotation terminology complicates cross-study comparison. These changes may be driven by clonal expansions of T cells to viral antigens or medications. Prior studies have explored cohorts of antiparkinson drug-naive patients using flow cytometry and determined there were no significant effects on peripheral adaptive immunity^60,61^. Further, most PD patients in the current study were taking antiparkinson medication and the effects of other medications were considered. Interestingly, the CD8+ KLRF1+ population has been related to CMV seropositivity status^29^. While we refrained from including individuals with active infections in our cohort, we lacked information on CMV exposure.

Using V(D)J immune repertoire sequencing, we measured clonal diversity by D50 index and Shannon’s entropy. We show a sex-specific decrease in TCR diversity in male donors that is only present in healthy blood and not in PD blood. This aligns with the known earlier decline in TCR diversity in males during healthy aging^62,63^ and highlights the lack of this sex-associated healthy phenomenon in the context of disease. This contrasts with prior work reporting a diagnosis-specific D50 decrease in PD blood^18^. Repertoire-based analyses are sensitive to skewed sampling which we address with a larger cohort, multiple diversity metrics, and cell-count adjustment. The present work involves more than 10 times the sample number analyzed and 3 times the number of cells sampled compared to previous investigations. These findings reveal intertwined effects of sex and diagnosis on the peripheral immune system and contribute an extensive resource to the field. These repertoire findings naturally lead to the question of what antigens these clonally expanded T cells may recognize.

Viral infection has been linked to neurological diseases such as Alzheimer’s^19,64^ and Parkinson’s,^65,66^ with the potential for cross reactive epitopes between a virus and endogenous proteins. Our GLIPH analysis identified potential epitope annotations for TCRs using CDR3b motifs and HLA sharing, revealing that clonal expansions in PD-and prodromal-enriched groups involve different cell types than those in control blood. Since Sulzer and colleagues found that only approximately 40% of the PD participants were reactive to PD epitopes^25^, PD donors in these motif groups may be responsive in follow up alpha-synuclein stimulation studies. However, this approach is limited by reliance on viral-dominated reference databases and undersampling of HLA alleles. Definitive evidence for PD-specific antigen specificity will require stimulation experiments accounting for HLA context.

Together, these findings highlight alterations in blood immune cell type composition and T cell clonal characteristics that may synergize to contribute to inflammation in PD. Simultaneous increases in naive CD8+ T cells with decreased cytotoxic CD8+ T cell subtypes and TCR diversity, particularly in males, sheds insight into a potential sexual dimorphic underlying PD immune risk profile. Our study is the most extensive paired RNA and V(D)J scRNA sequencing focused on Parkinson’s disease and prodromal RBD cases. While scRNA-seq technology is excellent, such studies are limited by the necessity to balance the benefits of increased cell count and total donor number. As such, the per donor sampling is still limited and cannot completely capture all possible blood cell types and the complete immune repertoire diversity. Changes in T-cell subtypes should be confirmed with follow up targeted studies before expanding such results to clinical utility for biomarkers. Combining this data with other large scale studies would help even more completely assess lymphocyte diversity. As our understanding of the role of T cells evolves, this dataset will remain a powerful and useful resource to more deeply explore the peripheral immune system in PD.

## Methods

### Clinical centers and recruitment strategies

Participants were enrolled at The Marlene and Paolo Fresco Institute for Parkinson’s and Movement Disorders at the New York University Langone Health (New York) and The Mount Sinai-Beth Israel Movement Disorders Clinic at the Mount Sinai Hospital (New York). Each Institution’s Institutional Review Board (IRB) approved the study protocol and the related procedures for subject recruitment, as well as the data and sample collection plan. Subjects were enrolled only upon signing IRB-approved informed consent. Patients were diagnosed by neurology-board-certified movement disorders specialists. PD symptoms and disease stage were assessed using the Movement Disorder Society Unified Parkinson Disease Rating Scale (MDS-UPDRS), and Hoehn and Yahr (HY) scale, while cognitive profiles were evaluated with the Montreal Cognitive Assessment (MoCA) scale. Additional information such as family history, clinical history (recent infection and vaccination), medication history and the presence of non-motor features typically associated with PD were collected at the time of recruitment. The inclusion criteria for diagnoses were as follows: Prodromal are subjects without PD diagnosis, but with clinical risk factors of PD such as RBD (based on self-reported clinical symptoms or polysomnography); iPD are patients diagnosed with PD and without monogenic risk genes or other known genetic risk factors. Genetic manifesting carriers have a PD diagnosis and *GBA1* (N370S) / *LRRK2* (G2019S) mutations while non-manifesting carriers only have said mutations; Controls are age-matched subjects that do not have PD diagnosis in their personal or family history. Participants were recruited with concerted efforts to collect RBD prodromal and genetic carriers of Parkinson’s disease risk mutations. Our 176-participant cohort is composed of 51 sex-balanced and age-matched controls (∼45-70 years old), 20 prodromal patients with REM sleep behavior disorder (RBD), 52 idiopathic PD (iPD), and 39 genetic PD having either *GBA1* or *LRRK2* mutations, 3 genetic PD with both *GBA1* and *LRRK2* mutations and 11 non-manifesting genetic carriers of those same mutations.

### Blood collection and PBMCs isolation

A maximum of 30 mL of blood was collected in Vacutainer blood collection tubes with acid citrate dextrose (ACD) (BD Biosciences). Fresh blood was delivered to the Raj laboratory and processed within 2-3 hours. First, 3 mL of blood was centrifuged at 1,500g for 15 mins, and aliquots of whole blood were stored at -80°C for genotyping. Subsequently, remaining blood was diluted in 2-fold PBS (Gibco) and peripheral blood mononuclear cells (PBMCs) were isolated using SepMate tubes (StemCell Technologies) filled with 15 mL of Ficoll-Paque PLUS (GE Healthcare) through a 15 mins centrifugation at 1,200g. After washing with PBS, PBMCs counts and viability were assessed using Countess II Automated Cell counter (Thermo Fisher). PBMCs were cryopreserved in 90% FBS (Germini) +10% DMSO (Sigma Aldrich) at a concentration of 10 million cells/mL in Nalgene cryogenic vials (ThermoScientific). Vials were placed in NalGene CryoFreezing containers at -80°C during 24-72 hours, and subsequently placed in liquid nitrogen for long-term storage.

### Single-cell RNA-seq data generation and processing

Cryopreserved PBMCs were thawed and restored under standardized conditions. Following thawing, cell yield, purity, and viability were assessed. Single-cell RNA sequencing (scRNA-seq) libraries were generated *in-house* using the 10x Genomics 5’ v2 chemistry, multiplexing four samples (loading ∼49,500 cells total) per run, allowing us to capture on average ∼5,500 cells per donor prior to QC. Separate libraries were prepared for gene expression (GEX) and T-cell receptor (TCR) V(D)J profiling. All libraries underwent quality control using the Agilent TapeStation and were sequenced by Azenta Inc. on a NovaSeq 6000 platform, targeting ∼20,000 reads per cell for GEX and ∼5,000 reads per cell for V(D)J. On average, we captured 23,000 cells per run with around a read depth of 28,000 for GEX (**Supplementary Table 2**). Following sequencing, raw data were demultiplexed and returned to our lab for downstream analysis.

### Genotyping

Participants were genotyped using DNA extracted from 1 mL whole blood aliquots. We used the QIAamp DNA Blood Midi kit (Qiagen) and followed the manufacturer’s instructions. DNA quality and concentration was assessed using a NanoDrop. Samples were genotyped using the Illumina Infinium Global Screening Array (GSA), which contains a genome-wide backbone of 642,824 common variants plus custom disease SNP content (∼60,000 SNPs). We performed HLA imputation using SNP genotypes to obtain two-field HLA alleles which were used in the viral reactivity analysis. Additionally, we performed targeted genotyping for specific regions associated with neurodegenerative diseases (*LRRK2*, *GBA1* and *APOE*). *LRRK2* and *GBA1* genotyping was performed by Dr. William Nichols’ laboratory at the Cincinnati Children’s Hospital. SNP genotyping was performed for the G2019S variant in *LRRK2* and the 11 most common variants in *GBA1* (84GG, IVS2+1, E326K, T369M, N370S, V394L, D409G, L444P, A456P, RecNcil, R496H). We included patients with only the most common N370S *GBA1* mutation. The three major *APOE* isoforms (APOE 2, APOE 3, APOE 4) were assessed in the laboratory using TaqMan assays for both rs429358 (C 3084793_20) and rs7412 (C 904973_10) from Thermo Scientific following manufacturer’s instructions. 10 ng of DNA were added to the SNP reaction mix in a 96-well plate. Fluorescence reading of the TaqMan assays was performed using QuantStudio 7 Flex (Applied Biosystems).

### Ancestry estimation using genotypes

The following quality control metrics were applied to genotypes prior to ancestry estimation: minor allele frequency (MAF) >5%, SNP and samples call rate >95%, Hardy-Weinberg equilibrium (HWE) *P-value* >1×10-6. PLINK^67^ was utilized to identify and remove duplicated and genetically related samples using pairwise IBD (identity-by-descent) estimation (PLINK PI_HAT values 0.99-1). Somalier v.0.2.19^68^ was used to estimate genetic relatedness and ancestry. Genetic ancestry was confirmed through principal component analysis and comparing multidimensional scaling (MDS) of the values of the study cohort with data from the Phase 3 of 1000 Genome Project samples^69^. For the Ashkenazi Jewish (AJ) only, analyses were repeated using an Ashkenazi Jewish reference panel^70,71^.

### Primary scRNA-seq data processing

Raw sequencing gene expression (GEX) and immune repertoire (V(D)J) reads from Illumina NovaSeq Sequencer were aligned to the GRCh38 genome and V(D)J reference, respectively, using Cell Ranger v.8.0.1 *count* and *vdj* pipelines. Count matrices were incorporated in R as Seurat objects for downstream analysis using Seurat v.4.4.0^72^.

### Quality control and data processing

Samples that were processed in the same 10X run and share technical variation (4 donors per run) underwent standard pre-processing steps including the removal of cells with low and high gene count (excluding cells with <200 genes and > 3 standard deviations from the mean number of expressed genes) and high mitochondrial percentage (>10%), log normalization and scaling (**Figure S1**). SoupX v.1.6.2^73^ was used to estimate and remove ambient RNA. Donors were demultiplexed based on genotypes using Demuxlet^74^ (homotypic doublets) and additional heterotypic doublets were identified using DoubletFinder v.2.0.3^75^. Cells identified as doublets or negatives by either method were removed. On average across the data, there was a 71% recovery rate of singlets per run resulting in around 4,000 cells per donor (**Supplementary Table 2**). After demultiplexing, objects from all sequencing runs were merged then integrated into one Seurat object to streamline analyses and account for batch effects using Harmony^76^. Before moving forward with integrative analysis, 8 donors were removed because they were genetically identical to other donors in the cohort or could not be demultiplexed based on their genotype. Additional QC steps were performed on the integrated log-transformed counts prior to cell type-specific analyses separately. After QC, we maintained transcriptome data for 168 unique donors and over 630k cells.

### Broad cell type annotation

Unsupervised clustering was used to identify major cell types in the data. Dimensionality reduction using principal component analysis (PCA) was performed to reduce the feature space before clustering. Harmony was used to perform batch correction using the PCA reduction. Nearest neighbors were identified using the Harmony embeddings. We performed Louvain clustering using Seurat’s clustering functionalities on the first 36 Harmony embeddings which comprises 95% of variance within the data and with a resolution of 0.4. This resulted in 26 numbered clusters. The harmonized coordinates were visualized in 2-dimensional space using a uniform manifold approximation and projection (UMAP). The clusters were broadly annotated as 18 different PBMC cell types based on known marker gene expression and corroborated using reference-based annotation in SingleR v.2.2.0^77^ with the Monaco Immune Reference^44^ (**Figure S5**). In particular, the reference annotation was used to assist in the identification of Intermediate Monocytes. Clusters that did not express known markers of PBMCs (**Supplementary Table 3**) or that are indicated to be low-quality or doublets were removed. Platelets and red blood cell contamination was identified and removed based on normalized expression greater than 1 of any of the following genes: *PPBP*, *GNG11*, *PF4* and *HBB*.

Two clusters were annotated as Low Quality Monocytes and Low Quality T Cells based on their canonical gene expression, higher percent mitochondrial genes (>7.5%) and lower ribosomal genes (<5%) compared to other cells in the dataset. Cells in these clusters were removed based on their low ribosomal count. It should be noted that these cells also had low expression of housekeeping genes (*GAPDH*, *ACTB*, *APOO*, *EIF3K*) and their respective canonical cell type markers compared to other clusters.

### T cell and NK cell subclustering and annotation

Cells broadly labeled as NK and T cells were subset and reprocessed in the same way as was described above. In order to identify subpopulations, we explored the expression levels of curated marker gene lists which were compiled across published literature^78,79^, the AIFI Immune Health Atlas and Azimuth (**Supplementary Table 3**). Small numbers of contaminated cells were removed. After higher resolution clustering, 28 different PBMC cell types were more-specifically annotated.

T cells were isolated from NK cells and were further annotated using Symphony v0.1.1^43^ with rheumatoid arthritis (RA) cohort reference^42^. T cells were first separated from NK cells based on cell type clustering and confirmed by mapping the V(D)J gene library to our Seurat object. Data was projected onto the Symphony reference using the mapQuery and knnPredict functions to generate confidence scores for reference clusters’ mapping. The most likely cluster identities were cross-referenced with prior marker-based annotations to aid in generating final T cell cluster annotations (**Figure S5**). After higher resolution clustering and convergent methods for annotation, 18 different T-cell types were more-specifically identified.

### Cell-type compositional analysis

Cell type compositional changes were assessed using two complementary approaches: Covarying Neighbor Analysis (CNA; rcna implementation 0.0.99^80^), which identifies transcriptional neighborhoods enriched across disease stages, and linear regression model (crumblr v.0.99.6^34^) to confirm findings and explore interaction effects (**Supplementary Table 5**). CNA analysis was performed to test the associations of Diagnosis, Age, and Sex with cell composition. In each model, one variable served as the primary predictor while the remaining ones were included as covariates. Sequencing batch was used as the batch variable in all of the analyses. Crumblr analysis was performed using the counts for each of the discrete cell types and determining their association within a single precision-weighted linear (mixed) model framework. The normal approximation of transformed count data from a Dirichlet-multinomial model allows use of standard workflows to analyze count ratio data while modeling heteroskedasticity^34^. A main effects model was used including Age, Sex, Diagnosis status (case-control), *LRRK2* G2019S mutation status, *GBA1* N370S mutation status and Ancestry (AJ or non AJ). An interaction effects model was used to query the way sex and diagnosis jointly impact cell type composition. The same covariates were used with the addition of an interaction term (Diagnosis*Sex). Corrected fold changes were reported and significant changes were determined using a p-value cutoff of 0.05. Adjusted p-values were calculated using the p.adjust in the stats R package.

### Differential gene expression analysis

We performed differential expression analysis focused on comparing the idiopathic PD cases and the genetic PD cases separately. We compared the following pairwise groups: iPD vs. Control, iPD vs. Prodromal, Prodromal vs. Control and GBA1-PD vs. iPD, GBA1-PD vs. LRRK2-PD, GBA1-PD vs. Control. Differentially expressed genes were identified using a pseudobulk approach using DESeq2 v.1.50.2^81^. Before running DESeq2, single cells were aggregated to pseudobulk at the cell type and sample level using the filterByExpr function from the EdgeR v.4.8.0 package^82^. For each cell type, outlier samples were removed if they had a z-score > 3 in PCA analysis of the variance stabilizing transformed (VST) pseudobulked expression data (**Figure S4**). Using this threshold, no more than 3 samples were removed as outliers for each cell type and comparison. The design matrix included diagnosis, sequencing batch, sex and age. Multiple testing correction was performed with the Benjamini-Hochberg (BH) procedure to control the false discovery rate (FDR). Genes were considered significantly deregulated when they showed an absolute log2 fold-change greater than 0.5 and an adjusted p-value less than 0.05 (**Supplementary Table 6**).

### Pathway enrichment analysis

Gene set enrichment analysis (GSEA) was performed on the DESeq2 differential expression results using clusterProfiler (v.4.10.1)^83^. Genes were ranked by signed significance from DESeq2 output, -log10(p-value)*sign(log2FoldChange), which represents the direction of effect and the significance. Pathways meeting an adjusted p-value threshold of 0.05 (method = “BH”) were kept. A data frame containing term ID, pathway or category description, GeneRatio, and adjusted p-value was used (**Supplementary Table 7**).

### Immune repertoire analysis

The Cell Ranger *vdj* pipeline output file filtered_contig _annotations.csv was used to identify TCR sequences obtained for each cell barcode. V(D)J reads were aggregated and read into scRepertoire v.2.4.0^84^, specifying a single donor to allow for the characterization of TCRs to be uniform. Individual donors were matched with their corresponding cells based on cell barcodes prior to clonotype quantification. Clonotypes were defined based on identical CDR3 amino acid sequences in both alpha and beta chains. We also alternatively defined clones using CDR3 nucleotide sequence and the strict approach using both CDR3 nucleotide sequences and gene usage. However, we focused on the CDR3 amino acid sequence similarity defined clonotypes because this emphasizes epitope recognition. scRepertoire was used to assess clonotype distribution, clonal diversity and clonal expansion (**Supplementary Table 10**). Clonotypes were added as metadata to our Seurat object using an *in-house* function, identifying donor origin by matching the cell barcodes to the already demultiplexed gene expression data. Over 337k cells had associated TCR information. Public clonotypes were defined as those coming from different donors and containing identical CDR3 nucleotide sequences and V and J genes.

TCR repertoire clonal diversity was measured using both D50 values and Shannon’s entropy. Shannon’s entropy was calculated using scRepertoire’s clonalDiversity function with the default 100 iteration bootstrapping to make a fair comparison between groups of different sizes. The D50 values were determined as the proportion of dominant TCRβ CDR3 clones in each sample that cumulatively account for at least 50% of the total TCRβ sequences^85^. The calculation formula was as follows:

1. The number of total TCRβ sequences in a sample is N and the number of all distinct TCRβ CDR3 amino acid sequence types is C.
2. The functional TCRβ sequences corresponding to each unique CDR3 sequence were defined as N_1_, N_2_, …, N_C_, and sorted in descending order of abundance where N_1_ ≥ N_2_ ≥ … ≥ N_C_ − 1 ≥ N_C_.
3. When (N_1_ + N_2_ + … + N_H_ − 1) ≤ 0.5 × N and (N_1_ + N_2_ + … + N_H_) ≥ 0.5 × N, H is the number of unique clones used when the cumulative sum approaches 50% of the total TCR sequences in a sample. D50 represents the ratio of H to C.

GLIPH2 irtools v.0.01^86^ was used to predict viral reactivity analysis or antigen specificity. Predictions are based on CDR3 amino acid matches between beta chains in the cohort versus public databases. A GLIPH2 TCR input file was created from our 5’ V(D)J sequencing information (CDR3β, TRBV and TRBJ genes, CDR3α and frequency) and only cells with paired gene expression information were used. A corresponding HLA input file was made using imputed HLA genotypes from our cohort. Any missing input information was indicated with an NA value in accordance with the GLIPH guidelines. The initial GLIPH2 run with only our cohort was used to establish T-cell specificity groups. To further attempt to annotate specificity groups a combined GLIPH analysis was performed using both our cohort and publicly available CDR3 sequences from VDJdb^87^, McPAS-TCR^88^, iedb^89^ and PIRD^90^, obtained from the Trex package^84^. Annotations across databases were simplified favoring the more common antigens as primary predicted specificity. Each entry or combination of CDR3β and TRB genes was treated as a donor and frequency values were set to the number of databases the clonotype was found in to meet the requirements of GLIPH2 input. Results were filtered to focus on groups with patterns matching more than three amino acids and having membership from more than three donors (**Supplementary Table 11**). For antigen specificity comparison of the combined GLIPH2 run, results were filtered to retain GLIPH groups enriched in PD and prodromal donors, focusing on TCRs considered clonal (with a frequency greater than 2) in PD or prodromal donors and further filtered for stringency with a Fisher score <0.05 (**Supplementary Table 12**).

### Statistics and reproducibility

Significance was measured using marginal p-values or adjusted p-values which were corrected for multiple testing using Benjamini-Hochberg (BH). Each analysis indicates whether a more stringent or lenient significance metric was used and which threshold value was used.

### Data Availability

All summary statistics are provided in the Supplementary Tables. Differential expression results for all cell types and subtypes are included in Supplementary Table 6. Raw scRNA-seq data and processed read counts will be made available via CELLxGENE upon publication. Genotype data will be deposited in dbGaP under accession ID phs002400.v1.p1.

### Code Availability

The code used for the primary analysis is available on GitHub at: https://github.com/RajLabMSSM/sc_PBMC_PD. Any additional code used for analysis is available upon request from the corresponding author.

## Acknowledgments

We thank the study participants for their blood donations. T.R. is supported by U.S. National Institutes of Health (NIH) grants NINDS R01-NS116006, R01-NS133742, U01-NS107016 and U54-NS123743, and NIA R21-AG063130, R01-AG054005, U01-AG068880, R01-AG065926, R56-AG055824 and P30-AG066514, and by funding from Calico Life Sciences and Denali Therapeutics. R.S-P. is supported by NINDS U01-NS107016 and U01-NS094148, the Bonnie and Tom Strauss Chair, and the Bigglesworth Family Foundation. M.R. is supported by NINDS F31-NS134319 and a Parkinson’s Foundation Visiting Scholars Award. We thank Dr. Soumya Raychaudhuri and his lab members Dr. Jose Alquicira Hernandez and Dr. Yu Zhao for their support and feedback on this analysis during Mikaela’s Parkinson’s Foundation Visiting Scholars Award. O.N.M. and D.M. are supported by Silverstein Foundation Fellowships. Additional support was provided by the Office of Research Infrastructure Programs of the NIH under award numbers S10OD018522 and S10OD026880. The content is solely the responsibility of the authors and does not necessarily represent the official views of the Funders.

This work was supported in part through the computational resources and staff expertise provided by Scientific Computing at the Icahn School of Medicine at Mount Sinai and supported by the Clinical and Translational Science Awards (CTSA) grant UL1TR004419 from the National Center for Advancing Translational Sciences. Research reported in this paper was supported by the Office of Research Infrastructure of the National Institutes of Health under award number S10OD026880 and S10OD030463.

## Author Contributions

T.R., O.N.M. and M.R. conceived and designed the study. O.N.M., E.M., D.M., C.P.M., A.A., T.K. and C.A. processed samples and generated the data. M.R analyzed the data and performed statistical analyses with assistance from T.N. and B.J. with additional supervision from T.R. and O.N.M. M.R., O.N.M., A.T., G.R., R.S-P. and T.R. interpreted the results. M.R. and O.N.M. wrote the manuscript and M.R., O.N.M. and T.R. edited the manuscript. All the authors have read and approved the final manuscript. T.R. supervised the study. The funders had no role in study design, data collection and analysis, decision to publish, or preparation of the manuscript.

