## Supplementary Figures for "Peripheral Immune Alterations and T Cell Clonal Dynamics Across the Parkinson’s Disease Spectrum"

**Table of Contents**

**Supplementary Figures** **2**
Supplemental Figure 1. scRNA-seq quality control (QC) steps and metrics. 2
Supplemental Figure 2. Marker gene expression for PBMC populations. 3
Supplemental Figure 3. Comparison of cell type composition methods CNA and Crumblr and various models. 4
Supplemental Figure 4. Differential expression correlation between various models and methods. 6
Supplemental Figure 5. T cell annotations informed by projection onto Symphony reference. 8
Supplemental Figure 6. T cell subtype composition, method comparison and boxplot of cell counts. 9

**Supplementary Figures**

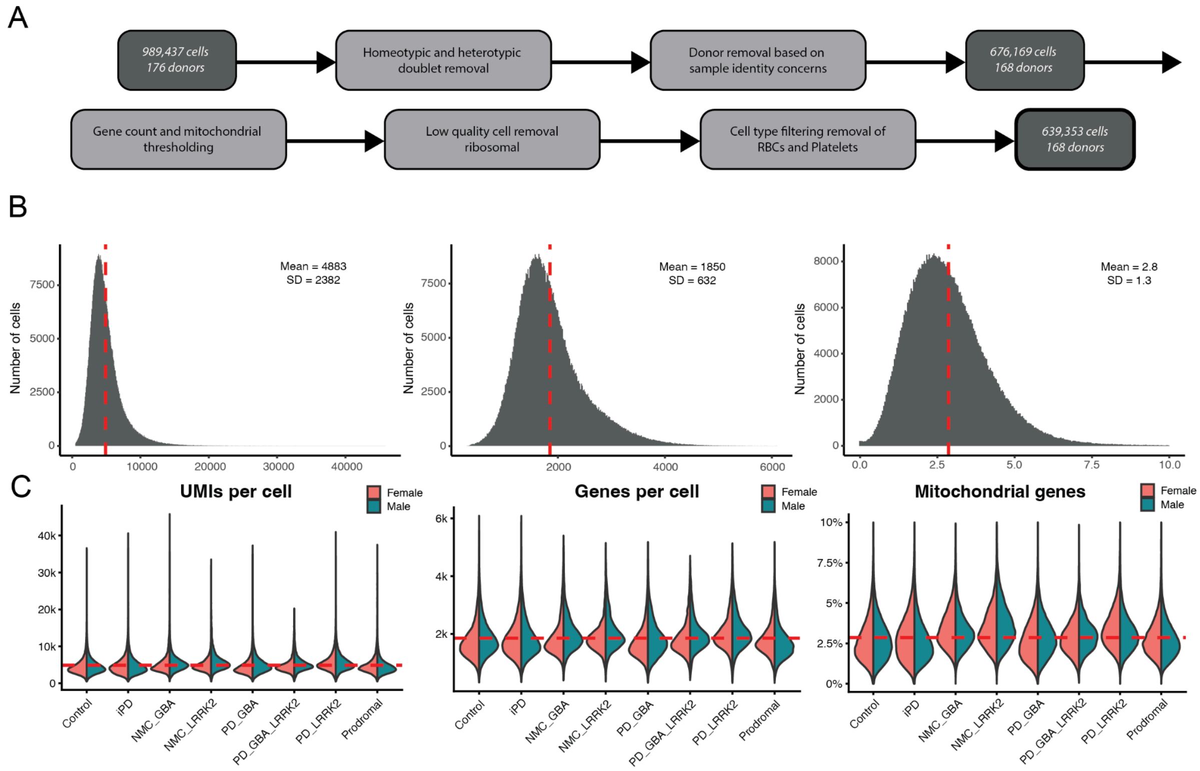


**Supplemental Figure 1. scRNA-seq quality control (QC) steps and metrics.**
**A)** Schematic outlining of QC steps and the amount of cells remaining after each step. **B)** Summary metrics after QC including the number of UMIs per cell, genes per cell and percent mitochondrial genes. Filters were applied to exclude cells with high or low overall gene expression per cell as well as those with greater than 10% mitochondrial genes. **C)** Distribution of the same QC measures shown in B, split by diagnosis and sex. In all panels, a dotted red line indicates the mean value.

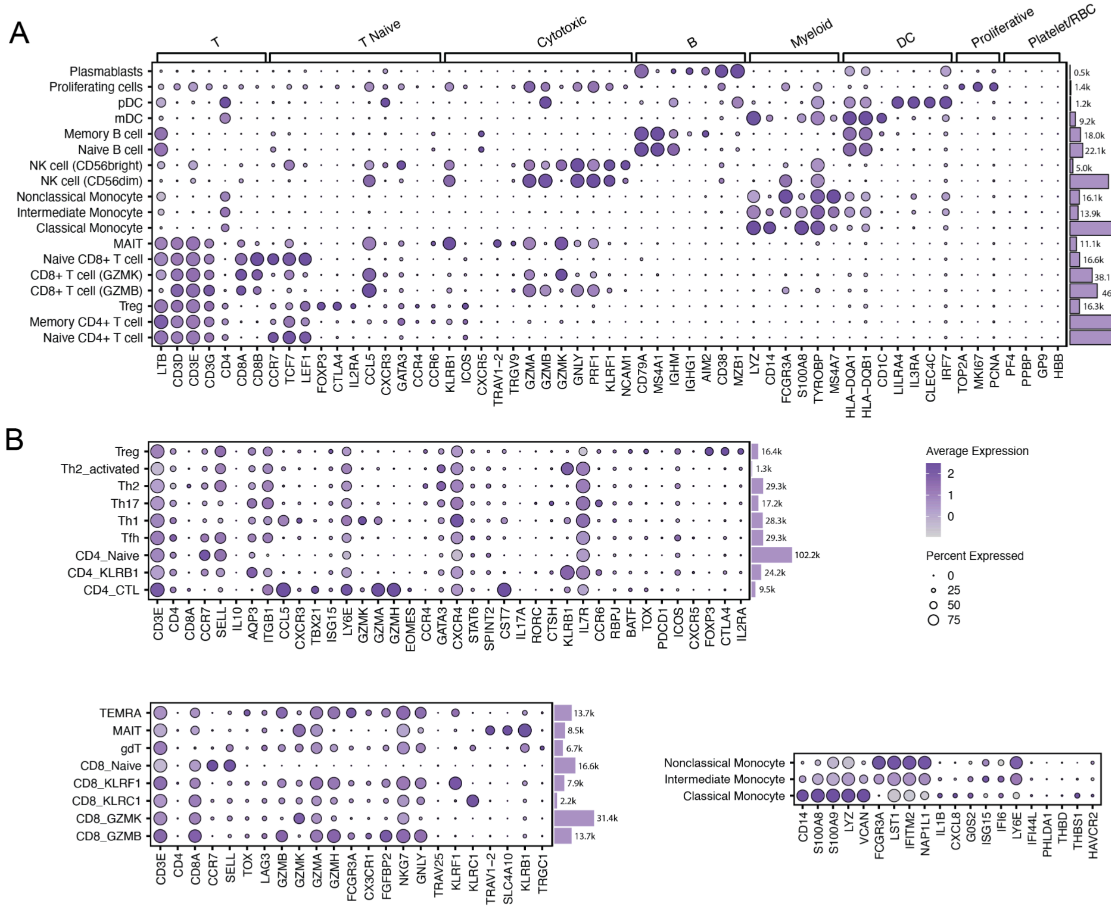


**Supplemental Figure 2. Marker gene expression for PBMC populations.**

Dot plots showing average expression of canonical expression markers (*x-axis*) characterizing each cell type (*y-axis*) for the **A)** general PBMC population and **B)** more specifically for subtypes. Dot size corresponds to the percentage of cells expressing the gene in each cluster and the color scale indicates the log normalized average expression of the gene in each cluster. Bars on right indicate the number of cells for each cell type.

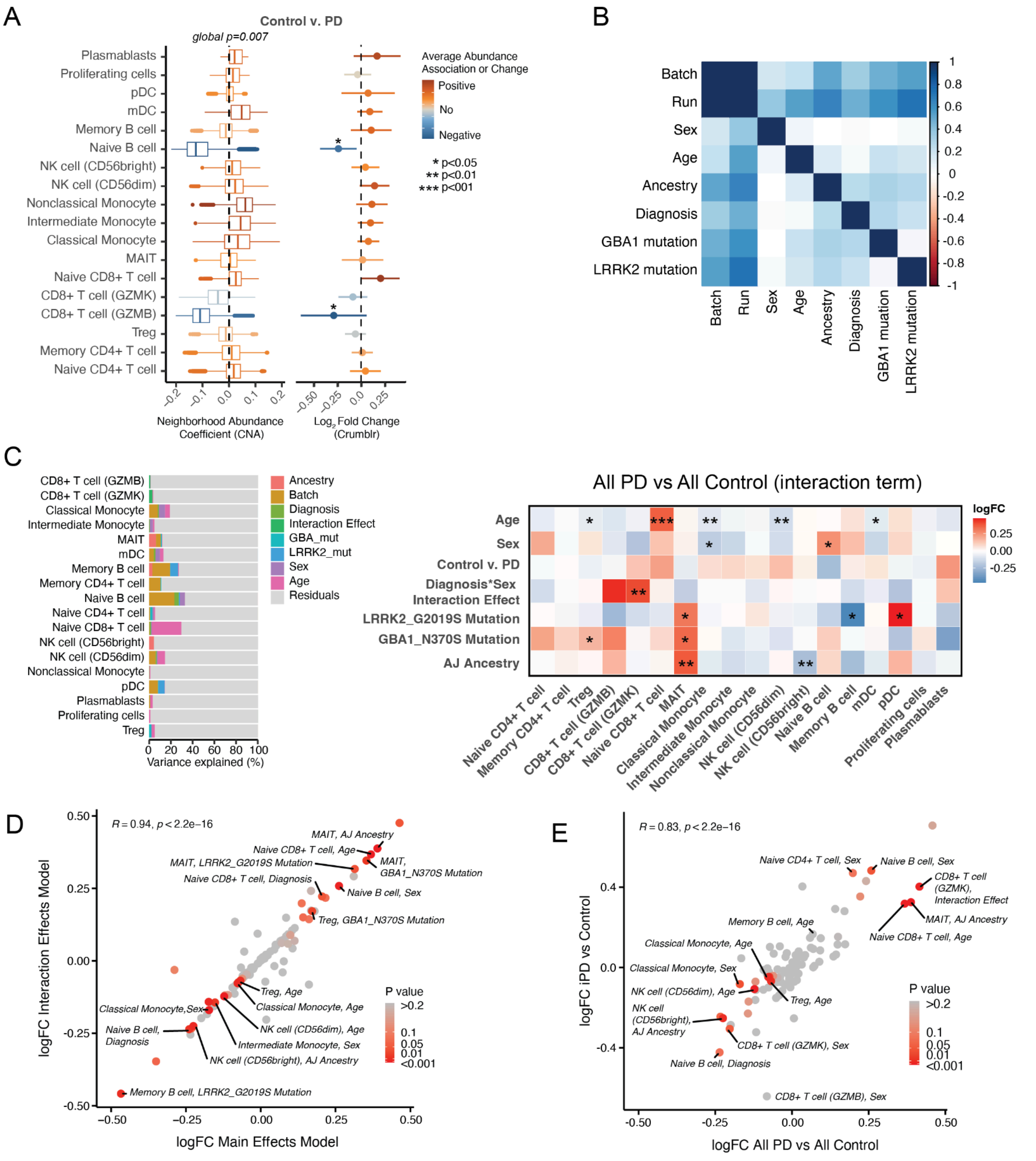


**Supplemental Figure 3. Comparison of cell type composition methods CNA and Crumblr and various models.**

**A)** Boxplot of CNA's Neighborhood Abundance Coefficients and Crumblr's Log2 Fold Change values for the Diagnosis comparison (Control vs PD) illustrating concordance between methods. Red color corresponds to positive values which indicates higher abundance association or change in PD, while blue corresponds to negative values which indicates higher abundance association or change in Control. Global significance in CNA for these neighborhood wide changes is p=0.007 and significant Crumblr changes are indicated by asterisks. **B)** Correlation matrix of metadata values. Colored squares indicate the strength and direction of the pairwise correlations, ranging from -1 (dark red) to +1 (dark blue) as indicated by the color scale bar. **C)** Variance partition plot and Crumblr results with an interaction term. **D)** Correlation between Fold Change effect sizes using a Main Effects model and Interaction Effects model. **E)** Correlation between Interaction models using the entire cohort (Control and NMC vs. iPD and gPD) versus clean cases (Control vs iPD).

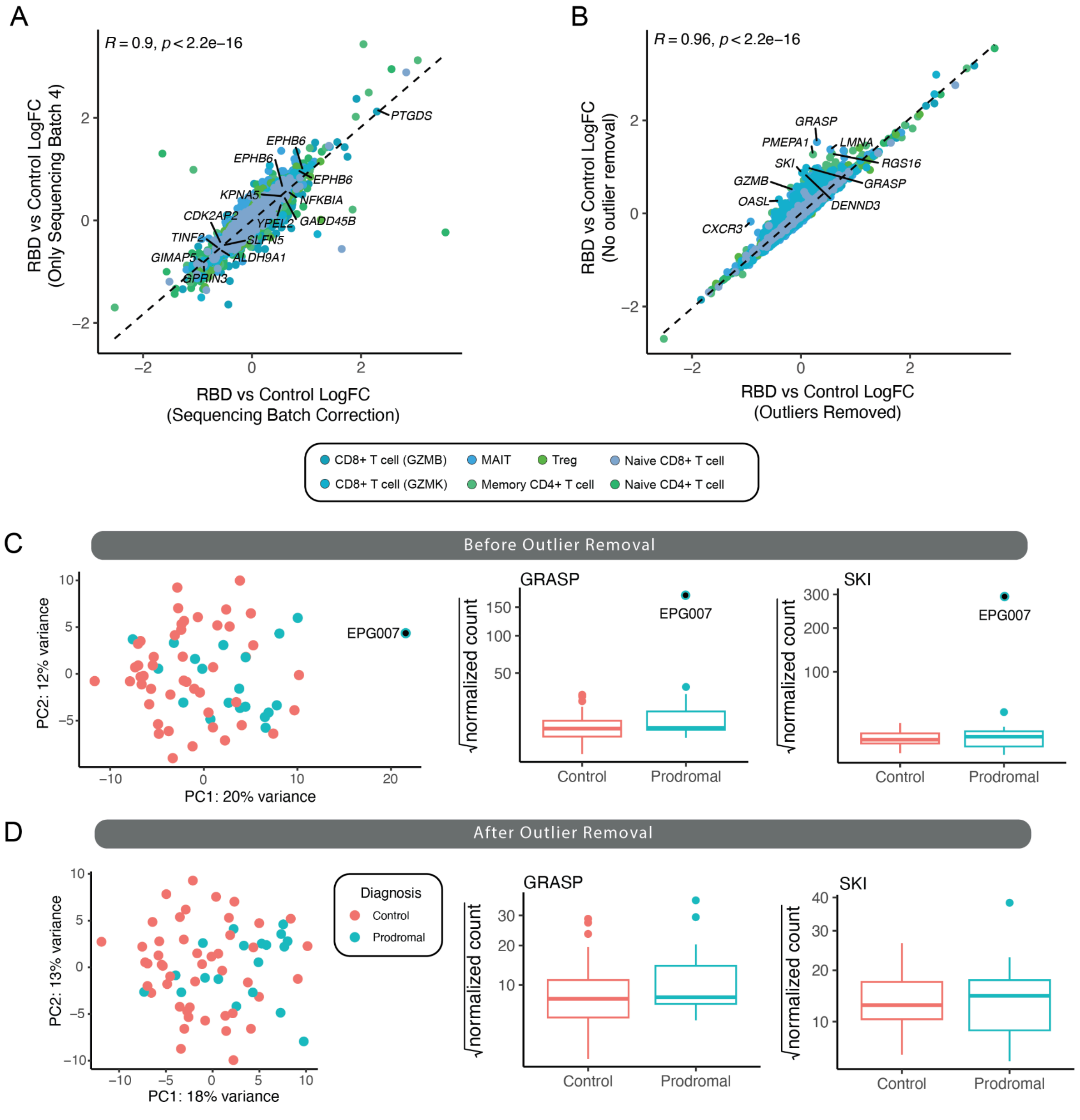


**Supplemental Figure 4. Differential expression correlation between various models and methods.**

**A)** Correlation of logFC values from pseudobulked DESeq2 analysis for Prodromal RBD and Control when only using samples in a single sequencing batch compared to using all sequencing batches with Batch correction incorporated in the model. Some of the most significant genes are labeled. **B)** Correlation of logFC values from pseudobulked DESeq2 analysis for RBD and Control comparing using or removing outliers. The top 10 most altered between the two analyses are labeled. **C-D)** Principle component analysis (PCA) of the normalized pseudobulked gene expression and square root of those counts for representative genes that are impacted by sample outlier shown including outlier sample (**C**) and with the outlier removed (**D**).

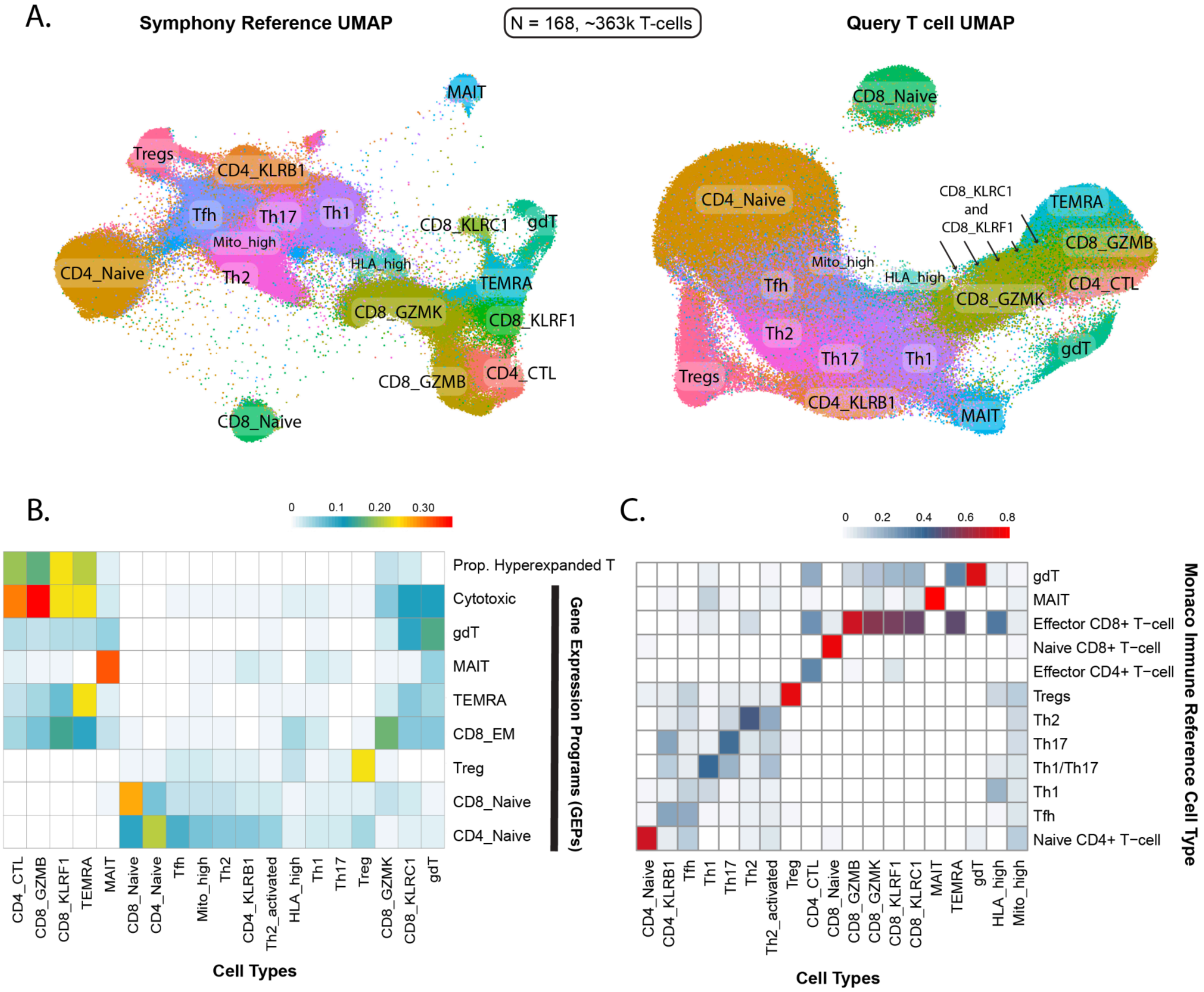


**Supplemental Figure 5. T cell annotations informed by projection onto Symphony reference.**

**A)** UMAP of T cell type annotations based on marker genes and projection onto a Symphony reference that contains a traditional 3' scRNA-seq and a 58-protein CITE-seq panel (left) and those same annotations projected back onto the T cell UMAP from this cohort. **B)** Proportion of hyperexpanded clones and mean gene expression program (GEP) usage for the most highly expressed GEPs in all T cell types. GEPs from Tcat can be used to help annotate cell types. The most cytotoxic cell types have the highest usage of Cytotoxic GEP and the most hyperexpanded clones. **C)** Cell type annotation comparison between final T cell subtypes and Monaco Immune Dataset.

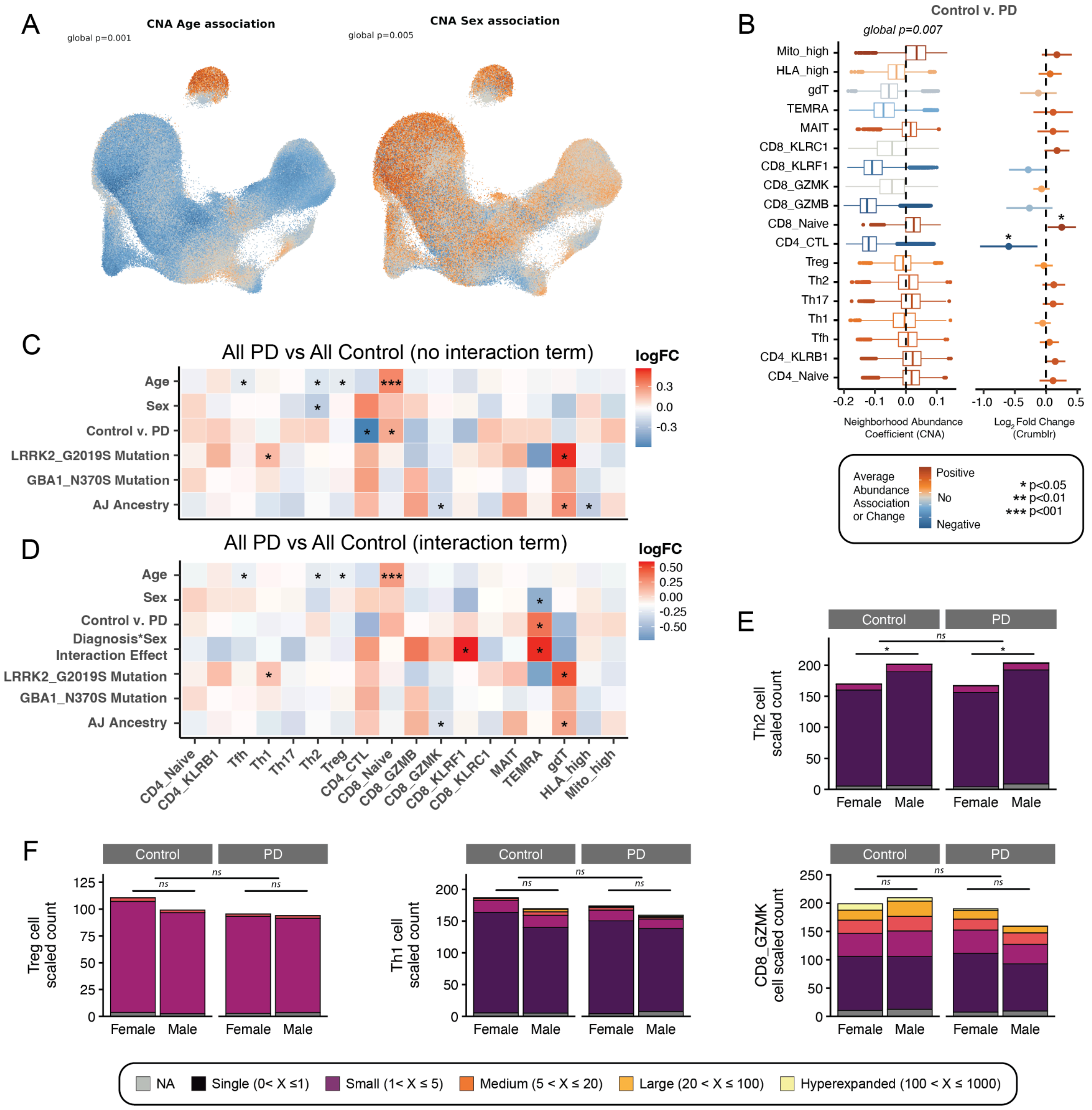


**Supplemental Figure 6. T cell subtype composition, method comparison and boxplot of cell counts.**

**A)** UMAP of T cells colored according to their neighborhood's abundance correlation to respective phenotypes, Age and Sex (from left to right). CNA global p-value is shown. **B)** Boxplot of CNA's Neighborhood Abundance Coefficients and Crumblr's Log2 Fold Change values for the Diagnosis comparison (Control vs PD) illustrating concordance between methods. Red color corresponds to positive values which indicates higher abundance association or change in PD, while blue corresponds to negative values which indicates higher abundance association or change in Control. Global significance in CNA for these neighborhood wide changes is p=0.007 and significant Crumblr changes are indicated by asterisks. **C-D)** Crumblr results without **(C)** and with **(D)** an interaction term modelling the effect of Diagnosis*Sex. **E-F)** Cell type counts for various T cell subtypes filled with proportion of clonal expansion. Counts were scaled by the total number of cells sampled per category. Th2 cells **(E)** showed a main effect with Sex. Tregs, Th1 and CD8+ (GZMK+) T cells **(F)** provide examples of cell type composition fluctuations without any significant effects for Diagnosis, Sex or the interaction of the two variables in our models **(C-D)**.
